# Menopausal vasomotor symptoms and subclinical atherosclerotic cardiovascular disease – a population-based study

**DOI:** 10.1101/2023.04.20.23288894

**Authors:** Sigrid Nilsson, Angelika Qvick, Moa Henriksson, Sofia Sederholm Lawesson, Anna-Clara Spetz Holm, Karin Leander

**Affiliations:** Department of Obstetrics and Gynecology in Linköping, and Department of Biomedical and Clinical Sciences, Linköping University, Linköping, Sweden; Unit of Cardiovascular and Nutritional Epidemiology, Institute of Environmental Medicine, Karolinska Institutet, Stockholm, Sweden; Department of Cardiology, and Department of Health, Medicine and Caring Sciences, Linkoping University, Linkoping, Sweden

**Author notes:** Corresponding author: Sigrid Nilsson Department of Obstetrics and Gynecology in Linköping, and Department of Biomedical and Clinical Sciences, Linköping University, Linköping 58185 Linköping, Sweden.

**Keywords:** Vasomotor symptoms, Menopause, Menopausal symptoms, Subclinical atherosclerosis, Cardiovascular disease.

## Abstract

**BACKGROUND:** Menopausal vasomotor symptoms (VMS) have been associated with subclinical and manifest atherosclerotic cardiovascular disease (ASCVD) but have not been studied in relation to image-detected coronary atherosclerosis. We assessed the association between VMS and subclinical ASCVD in peri- and postmenopausal women, considering a wide range of cardiovascular related risk factors that could potentially influence the relationship.

**METHODS:** This cross-sectional population-based study was conducted on a subset of participants from the Swedish CArdioPulmonary BioImage Study (SCAPIS), including women 50-65 years of age. The women underwent comprehensive cardiovascular assessments and completed an extensive questionnaire, which included questions about current and previous menopause-related symptoms. VMS was assessed on a 4-point scale and analyzed in relation to subclinical ASCVD, detected via coronary computed tomography angiography (CCTA), coronary artery calcium score (CACS) and carotid ultrasound using logistic regression analyses.

**RESULTS:** Of 2995 women included, 14.2% reported previous or on-going severe VMS (n=425), 18.1% moderate VMS (n=543), and 67.7% no or mild VMS (n=2027). Women who had ever experienced severe VMS, but not those with ever moderate VMS, had higher prevalence of CCTA-detected coronary atherosclerosis (34.1 vs 27.8%, p=0.017), but not segmental involvement score (SIS) >3 (4.5 vs 5.1%, p=0.332), CACS>100 (5.8 vs 6.8%, p=0.166) or any carotid plaque (47.6% vs 46.6%, p=249) than women with never or ever mild VMS. Using the same reference, ever severe but not moderate VMS was significantly associated with CCTA-detected coronary atherosclerosis, odds ratio (OR) after multivariable adjustment 1.33, 95% CI 1.02–1.72. This association was only present for durations of severe VMS of more than 5 years (multivariable adjusted OR 1.53 95% CI 1.09-2.14) or when the onset of severe VMS occurred before menopause (multivariable adjusted OR 1.60 95% CI 1.06-2.42). Additional adjustment for menopausal hormone therapy strengthened the associations whereas additional adjustment for physical activity did not. No significant association with SIS>3, CACS>100, nor with any carotid plaque was observed.

**CONCLUSIONS:** Previous or on-going severe but not moderate VMS were significantly associated with CCTA-detected coronary atherosclerosis, independent of a broad range of cardiovascular risk factors. No corresponding associations was observed for SIS>3, CACS>100 or carotid atherosclerosis.

## 1. Introduction

Atherosclerotic cardiovascular disease (ASCVD) is a major cause of mortality and morbidity in women (1), to which female reproductive factors appear to contribute (2, 3). To improve primary prevention strategies for ASCVD in women, there is a need for greater understanding of female specific risk factors that can be the focus of targeted preventive efforts (4).

Susceptibility to ASCVD in women increases considerably after menopause (5, 6), when estrogen levels decline sharply (7) and the prevalence of cardiovascular risk factors increases (8, 9). The decline in estrogen levels is often accompanied by climacteric symptoms (7), such as hot flushes and night sweats, known as vasomotor symptoms (VMS), affecting approximately 80-97% of women (10, 11) with varying frequency and severity (10, 12). The VMS duration is usually between 5 and 7 years (11, 13), but with an early onset the duration appears to be longer (10). There is an association between VMS and sleep-related problems, depressive symptoms, and a generally impaired quality of life (13, 14), especially among women with long VMS duration (13).

The most effective VMS treatment is menopausal hormone therapy (MHT) with either estrogen alone or in combination with gestagen. The prescription rate, however, has been relatively low for the past two decades, mainly due to cardiovascular safety concerns (15). Whereas endogenous estrogen appears to counteract the early atherosclerosis process (16), the menopausal transition is linked to progressive changes in tissue estrogen receptor characteristic (17). Consequently, the response to exogenously administered estrogen may differ depending on when treatment is initiated (18). Current guidelines support MHT for bothersome VMS in women within 10 years since menopause or ≤60 years of age at treatment initiation (19). Another proposed therapy for treatment of VMS is physical activity (20), but as the evidence is ambiguous it is not included in prevailing VMS treatment guidelines.

Women with VMS appear to have an unfavorable cardiovascular risk profile and previous observational studies have found associations with cardiovascular risk factors such as hypertension, hypercholesterolemia, and overweight/obesity (21). Studies on measures of subclinical ASCVD have yielded discrepant results. Associations with carotid intima media thickness (C-IMT) and aortic calcification have been demonstrated (21–24) but not with coronary artery calcium (CAC) (21). In prospective studies, associations between VMS and major adverse cardiac events (MACE) have been observed (25, 26). However, no distinct mechanism has yet been identified and it is still unclear whether VMS constitutes an independent risk factor for developing ASCVD.

To gain a better understanding of how VMS are associated with ASCVD, population-based studies are needed, allowing for comprehensive adjustments. Using subclinical outcome measures has the advantage of investigating associations with early-stage ASCVD when the opportunities for preventive interventions are greatest. Previous research addressing a possible association between VMS and subclinical ASCVD has been limited to the use of CAC and carotid ultrasound measurements as outcomes. The current study adds coronary computed tomography angiography (CCTA), a low dose imaging technique measuring coronary atherosclerosis with the ability to identify both the anatomical distribution and the characteristics of coronary atherosclerosis and plaques (27).

The current study aimed to assess associations between VMS and subclinical ASCVD detected through CCTA, CACS or carotid ultrasound, and to determine if potentially observed associations are modified by cardiovascular related risk factors.

## 2. Material & Methods

### 2.1. Study design and population

This is a sub-study of the Swedish CArdioPulmonary BioImage Study (SCAPIS) - a multicenter population-based observational study aimed at improving prediction and prevention of CVD (26). The SCAPIS protocol, described in detail elsewhere (26), was consistent for all centers and included collection of data through a comprehensive questionnaire, anthropometry, electrocardiogram (ECG), blood pressure, accelerometer measurements of physical activity, blood sampling, and extensive cardiovascular imaging.

Female study participants at two of the SCAPIS centers (Stockholm and Linköping) were invited to participate in the sub-study Survey on Women’s Health (SWH), requiring completion of an additional questionnaire including questions about menopausal status, age at menstrual cessation, menopausal symptoms including VMS, and the use of MHT and naturally derived compounds. The questionnaire was first tested in a small pilot study and some adjustments for greater clarity were made. Of the women invited to the SWH, 98% (n = 5043) agreed to participate. Among those, 578 women gave no information about menopausal status and were therefore not considered. The current study also has the following exclusion criteria: 1) being premenopausal (i.e., regular menses during the last year), 2) loss of menstruation due to other causes than natural menopause (hormonal treatments, surgical menopause, excessive exercising or dietary restrictions, pregnancy or breastfeeding), and 3) missing data on VMS.

The data collection took place between October 2015 and March 2018 in Stockholm and between October 2015 and June 2018 in Linköping. Data from SCAPIS were linked through each individual’s unique personal identification number to the National Patient Register and the National Prescribed Drug Register, maintained by the National Board of Health and Welfare, in order to obtain information about medical history.

### 2.2. Menopause and vasomotor symptoms

Menopausal status was defined as perimenopausal (<12 months amenorrhea), early postmenopausal (≥12-36 months amenorrhea), late postmenopausal (>36-72 months amenorrhea) or very late postmenopausal (>72 months amenorrhea).

The SWH asked whether the women had ongoing or previous VMS, and if so to what degree and with what duration. Information on age at the start of VMS was requested. In case of MHT, the instruction was to report VMS as they were before medication initiation. A four-point intensity scale was used, in accordance with the structure of “The Greene climacteric scale” (28), an established questionnaire to identify VMS: 1) never, 2) mild, 3) moderate, or 4) severe VMS. Herein, ongoing or previous exposure to VMS, regardless of intensity, are collectively referred to as ‘never’, ‘mild’, ‘moderate’ or ‘severe’ VMS. The questionnaire included a question on frequency of VMS in the last two weeks, with five predefined response options: 1) no symptoms, 2) occasionally, 3) a few times per week, 4) once a day, and 5) several times per day. Women also reported on whether they had other menopausal symptoms or ever used naturally derived herbal medicine as treatment (e.g., phytoestrogens) for VMS.

### 2.3. Coronary and carotid atherosclerosis measures

CCTA data were collected using multi-slice CT scanner (Siemens, Somatom Definition Flash, Siemens Healthineers, Erlangen, Germany). Identical software and hardware were used at both study sites. Details of image collection, processing, analyses, reconstruction, reading, and scoring are published elsewhere (29). Atherosclerosis was reported in 18 segments (30), with findings from the most clinically relevant segments (1 through 3, 5 through 7, 9, 11 through 13, and 17) mandatory to report. Coronary artery status was classified as no atherosclerosis, any atherosclerosis, nonsignificant (i.e., 1% to 49%) stenosis, significant (i.e., ≥50%) stenosis, missing segment or not assessable due to calcium blooming or technical failure. The coronary segmental involvement score (SIS) was calculated by adding the segments with present atherosclerosis and a score >3 indicates more widespread atherosclerosis.

The CAC images were acquired using ECG-gated non-contrast CT at 120 kV and CAC score (CACS) was calculated after summing the calcium content in each coronary artery according to Agaston (31). In regression analyses, CACS was dichotomized into 0-100 or >100 Agaston units.

The carotid arteries were assessed bilaterally for atherosclerotic plaques in the common carotid artery, bulb, and the internal carotid artery. Results were categorized as no, unilateral, or bilateral plaques. Siemens Acuson S2000 ultrasound scanner equipped with a 9L4 linear transducer (Siemens Healthineers, Erlangen, Germany) was used according to a standardized protocol. The definition of plaques is described in more detail elsewhere (32, 33).

The primary measures of subclinical ASCVD in the present study were 1) any coronary atherosclerosis, 2) SIS >3, 3) CACS >100, and 4) any carotid plaque.

### 2.4. Covariates

The procedures behind the physical examinations, anthropometry and the collection and analyzing of blood samples have previously been described (29). Based on questionnaire data, binary variables for educational level, country of birth, employment status, financial strain, quality of sleep, continuous stress, depressive symptoms, and specific medical diagnoses were created. Details about how all variables were created are given in supplement. The participants were categorized as current, previous, or never smokers. Among smokers, the sum of cigarette pack-years was calculated. Data on family history of myocardial infarction (MI) and stroke, respectively, and alcohol habits were obtained from the questionnaire and treated as binary variables (see supplement).

Accelerometry was used to measure physical activity and sedentary time. Study participants wore a tri-axial accelerometer (ActiGraph LCC, Pensacola) on their right hip during waking hours for seven consecutive days. The ActiLife v.6.13.3 software was used to set up, download and process the data (34). The processed data were registered as counts per minute (cpm), with higher cpm indicating more intense physical activity. Physical activity intensities were categorized into 1); “Sedentary”; 0 and 199 cpm 2) “Light physical activity”; 200-2689 cpm and 3) ‘Moderate to vigorous (MVPA)’; ≥2690 cpm, based on percentage of all wear time, in accordance with previous studies (34). The variables, MVPA and sedentary, were used descriptively for this study and MVPA was used in the regression models.

Women who reported use or past use of systemic prescription MHT, either in the form of oral pills, transdermal patches or transdermal gels and irrespective of indication, were considered as ever users of MHT. Subsequently, a categorical variable was created considering duration of use: 1) never user, 2) 1-4 years, and 3) ≥5 years. For sensitivity analyses, a subcategory of women who initiated MHT >6 years since menopause was created.

### 2.5. Statistical analysis

Mean and standard deviation (SD) or median with 25^th^ and 75^th^ percentiles were calculated for continuous variables as appropriate, depending on data being normally distributed or not. Data distribution also determined whether independent T-tests or Mann-Whitney U tests were used to assess statistical significance in comparisons across groups; the Chi^2^ test was used for categorical variables. Multivariate logistic regression models were performed to assess associations between VMS and subclinical ASCVD, with results presented as odds ratios (OR) with 95 % confidence intervals (CI). Women who never had experienced VMS and women with mild VMS were combined (herein referred to as the never/mild group) and served as the reference category in all analyses, in comparison with the moderate and severe groups, respectively. Women with a history of ischemic heart disease (definition given in supplement), percutaneous coronary intervention (PCI) and/or coronary artery bypass grafting (CABG) were excluded in analyses of associations between VMS and coronary atherosclerosis and CACS, and women with a history of ischemic stroke were excluded when analyzing associations with carotid plaques (Figure 1). Crude univariate models, and five multivariate models adjusted for different sets of variables were used. Model 1 included age at time of study inclusion and site. Model 2 included model 1 and a block of socioeconomic, demographic, lifestyle related and clinical cardiovascular risk factors: educational level, country of birth, menopausal status, systolic blood pressure, waist circumference, low density lipoprotein (LDL) cholesterol, triglycerides, diabetes mellitus, medically treated hyperlipidemia, medically treated hypertension, and smoking status. Model 3 included model 2 and a block of additional factors previously shown to be more prevalent among women with VMS and suggested to associate with ASCVD: continuous stress, quality of sleep, depression, and sleep apnea. Model 4 and Model 5 included model 3 and in addition MVPA and MHT, respectively, to explore any change in OR point estimates.

**Figure 1.**
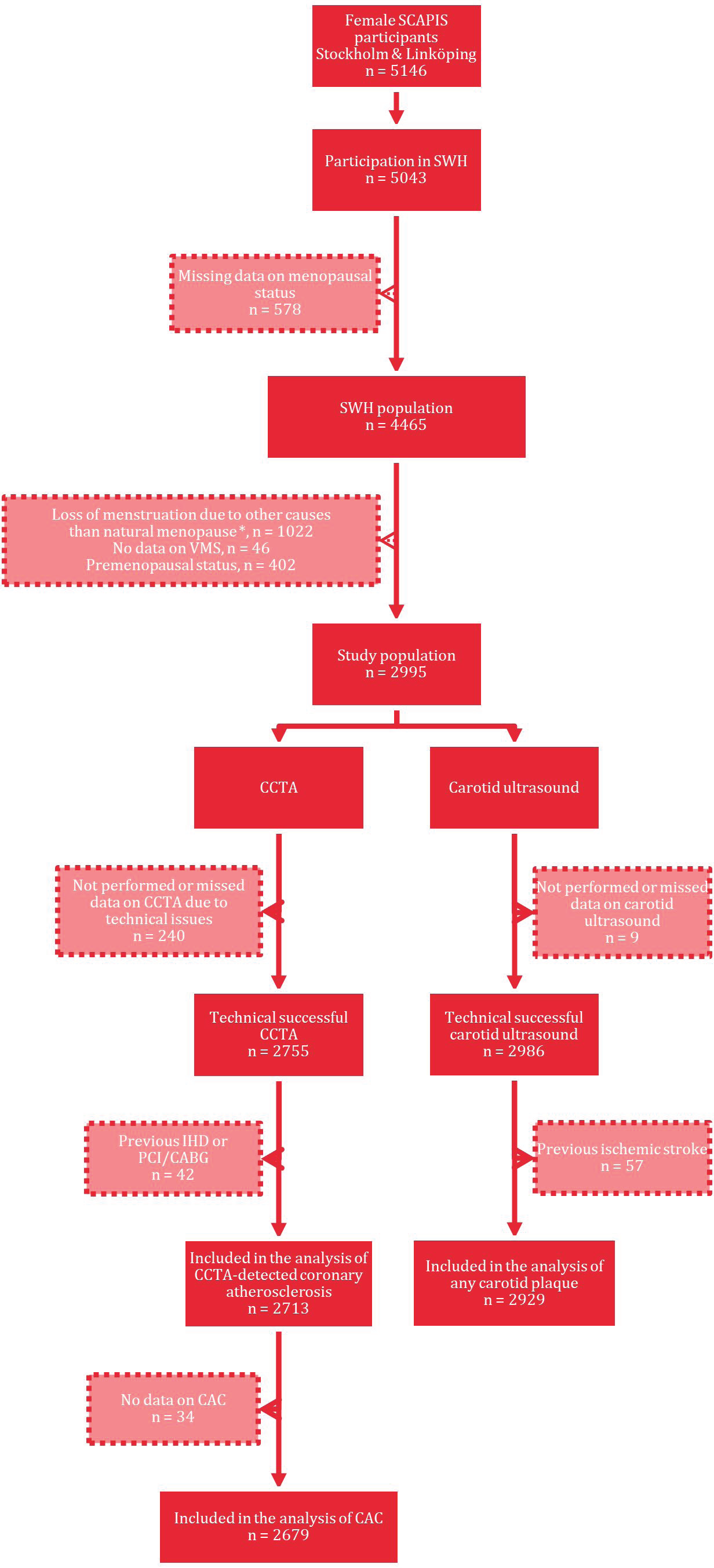
Flowchart for selection of the study population (n=2995) SCAPIS = Swedish CArdioPulmonary bioImage Study; ASCVD = Atherosclerotic cardiovascular disease; SWH = Study of Women’s health; VMS = Vasomotor symptoms; CCTA = Coronary computed tomography angiography; CAC = Coronary artery calcium; IHD = Ischemic heart disease; PCI = Percutaneous coronary intervention; CABG = Coronary artery bypass graft. * Hormonal treatments, surgical menopause, excessive exercising or dietary restrictions, pregnancy or breastfeeding.

To investigate whether duration of VMS had a bearing on the studied associations between VMS and subclinical ASCVD, we created a binary variable distinguishing longer and shorter duration. Since there is no established limit to make such distinction, we chose a cut-off corresponding to the median VMS duration in in the study sample. Then a variable was created with the following categories: 1) never/mild (reference), 2) moderate VMS ≤5 years, 3) moderate VMS >5 years, 4) severe VMS ≤5 years, and 5) severe VMS >5 years. Correspondingly, we investigated whether the timing of the start of the VMS had a bearing on the studied association between VMS and ASCVD. The variable created had the following categories: 1) never/mild VMS (reference), 2) moderate VMS staring before menopause, 3) moderate VMS starting after menopause, 4) severe VMS starting before menopause, and 5) severe VMS starting after menopause.

In supplementary analyses of VMS intensity in relation to coronary atherosclerosis, women with mild VMS served as the reference category, whereas women who had never experienced VMS constituted a separate category. Supplementary analyses were also performed to assess VMS in relation to CACS >0 as well as bilateral and unilateral carotid plaques. Finally, a sensitivity analysis was performed excluding women with late start (>6 years) of MHT. Self-reported physical activity was also considered instead of the objectively measured MVPA in a separate sensitivity analysis.

P-values <0.05 were considered to indicate statistical significance. All analyses were performed with SPSS v.28.0 (IBM, Portsmouth, UK).

## 3. Ethical considerations

This study was conducted in accordance with the Declaration of Helsinki, with oral and written informed consent before participation. The SCAPIS study as well as the SWH was approved by the Swedish Ethical Review Authority (reference numbers: 2010-228-31M, 2015-246/32, and 2020-03101). SN and KL had full access to all the data in the study and take responsibility for its integrity and the data analysis. The authors declare that all supporting data are available within the article.

## 4. Results

### 4.1 Descriptive characteristics

Among the randomly selected Swedish men and women invited to participate in SCAPIS, 30,154 individuals were recruited (50.3%). The participation rate was similar between sexes and ages (27). The mean age of the 4465 participants in the SWH population was 57.4 years; 82.6% were postmenopausal and 8.2% perimenopausal (characteristics presented in Supplementary Tables 1-3). After excluding participants according to the pre-specified criteria, 2995 women remained and constituted the study population (Figure 1). Of these, 43.3% reported mild, 18.1% moderate, and 14.2% severe VMS whereas 24.4% had never experienced VMS. Women in the severe group were more likely to be postmenopausal (93.4 vs 88.9%, p <0.01), and they experienced other menopausal symptoms (42.1 vs 24.3%, p<0.01) and used herbal medicines (e.g., phytoestrogens) (20.4 vs 5.5%, p<0.01) to a greater extent than women with never/mild VMS. Characteristics of the study population by VMS intensity are presented in Tables 1 and 2.

**Table 1.** General characteristics by VMS category of the study population, SCAPIS (n = 2995)

| Characteristics | Never/mild<br>n = 2027<br>67.7% | Moderate<br>n = 543<br>18.1% | Severe<br>n = 425<br>14.2% |
| --- | --- | --- | --- |
| **Age, mean (SD)<sup>†</sup>** | 58.3 (4.2) | 57.7 (4.3) | 58.4 (4.1) |
| **Born outside Sweden, n (%)<sup>\*</sup>** | 230 (11.5) | 53 (9.9) | 74 (17.7)<sup>#</sup> |
| **University degree, n (%)<sup>\*</sup>** | 1012 (50.7) | 285 (53.3) | 176 (42.1) |
| **Currently working, n (%)<sup>\*</sup>** | 1681 (83.9) | 462 (86.4) | 345 (82.3) |
| **Financial strain, n (%)<sup>\*</sup>** | 62 (3.1) | 18 (3.4) | 28 (6.7)<sup>#</sup> |
| **Smoking status, n (%)<sup>\*</sup>** |  |  |  |
| Past | 753 (38.1) | 217 (40.9) | 197 (48.0)<sup>#</sup> |
| Current | 209 (10.6) | 68 (12.8) | 55 (13.4) |
| **Pack years for cigarettes, median (25<sup>th</sup> – 75<sup>th</sup> percentile)<sup>‡</sup>** | 10.5 (4.5 – 19.5) | 11.8 (5.0 – 21.0) | 13.3 (5.5 – 21.0)<sup>§</sup> |
| **Medical history, n (%)<sup>\*</sup>** |  |  |  |
| Ischemic heart disease | 39 (1.9) | 5 (0.9) | 4 (0.9) |
| Ischemic stroke | 41 (2.0) | 11 (2.0) | 6 (1.4) |
| Hypertension | 353 (17.4) | 93 (17.1) | 80 (18.8) |
| Hyperlipidemia | 166 (8.2) | 31 (5.7) | 24 (5.6) |
| Diabetes mellitus | 78 (3.8) | 14 (2.6) | 16 (3.8) |
| Sleep apnea | 45 (2.2) | 8 (1.5) | 12 (2.8) |
| **Family history, n (%)<sup>\*</sup>** |  |  |  |
| Myocardial infarction | 144 (7.2) | 32 (6.0) | 32 (7.7) |
| Stroke | 126 (6.3) | 30 (5.6) | 40 (9.6)<sup>§</sup> |
| **Overweight (25-29.9 kg/m<sup>2</sup>), n (%)**<sup>*</sup> | 711 (35.1) | 224 (41.3) | 181 (42.6) |
| **Obese (≥30 kg/m<sup>2</sup>), n (%)**<sup>*</sup> | 379 (18.7) | 83 (15.3) | 79 (18.6) |
| **Waist circumference (cm), mean (SD)**<sup>†</sup> | 89.0 (12.9) | 88.8 (11.7) | 89.6 (11.7) |
| **Blood pressure (mmHg), mean (SD)**<sup>†</sup> |  |  |  |
| Systolic blood pressure<sup>†</sup> | 128 (18) | 128 (18) | 128 (19) |
| Systolic blood pressure >140, n (%)<sup>*</sup> | 490 (24.2) | 127 (23.4) | 102 (24.0) |
| Diastolic blood pressure<sup>†</sup> | 81 (11) | 81 (10) | 80 (11) |
| Diastolic blood pressure >90, n (%)<sup>*</sup> | 399 (19.7) | 98 (18.0) | 78 (18.4) |
| **Cardiometabolic biomarkers** |  |  |  |
| LDL cholesterol (mmol/L), mean (SD)<sup>†</sup> | 3.3 (1.0) | 3.3 (0.9) | 3.3 (1.0) |
| Triglycerides (mmol/L), mean (SD)<sup>†</sup> | 1.0 (5.3) | 1.0 (0.5) | 1.1 (0.7) |
| hsCRP (mg/L), median (25<sup>th</sup> – 75<sup>th</sup> percentile)<sup>‡</sup> | 0.9 (0.6 – 2.1) | 0.9 (0.6 – 1.9) | 1.1 (0.6 – 2.2) |
| HbA1c (mmol/mol) Mean (SD)<sup>†</sup> | 36.6 (5.9) | 36.3 (3.9) | 36.6 (5.7) |
| **Subjective experience of bad or very bad sleep, n (%)**<sup>*</sup> | 281 (14.2) | 106 (20.1) | 118 (28.9)<sup>#</sup> |
| **Physical activity, mean (SD)**<sup>†</sup> |  |  |  |
| Sedentary | 51.6 (10.0) | 51.4 (9.7) | 50.0 (9.7) |
| Moderate/vigorous activity | 6.7 (3.4) | 6.8 (3.3) | 6.5 (3.6) |
| **High alcohol consumption, n (%)**<sup>*</sup> | 160 (8.9) | 76 (15.4)<sup>#</sup> | 45 (12.3) |
| **Continuous stress last 1-5 years, n (%)**<sup>*</sup> | 463 (23.5) | 155 (29.2) | 133 (32.6)<sup>#</sup> |
| **Depression, n (%)**<sup>*</sup> | 405 (20.3) | 146 (27.3)<sup>#</sup> | 120 (29.2)<sup>#</sup> |

All statistical comparisons have the never/mild group as reference category. SCAPIS = Swedish CardioPulmonary bioImage Study; VMS = Vasomotor symptoms; SBP = Systolic blood pressure; DBP = Diastolic blood pressure; LDL = Low density lipoprotein; hsCRP = high sensitivity C-reactive protein. <sup>\*</sup> Chi2 test; <sup>†</sup> Independent T-test; <sup>‡</sup> Mann-Whitney U test; <sup>§</sup> p < 0.05; <sup>||</sup> p < 0.01; <sup>#</sup> p < 0.001.

**Table 2.**
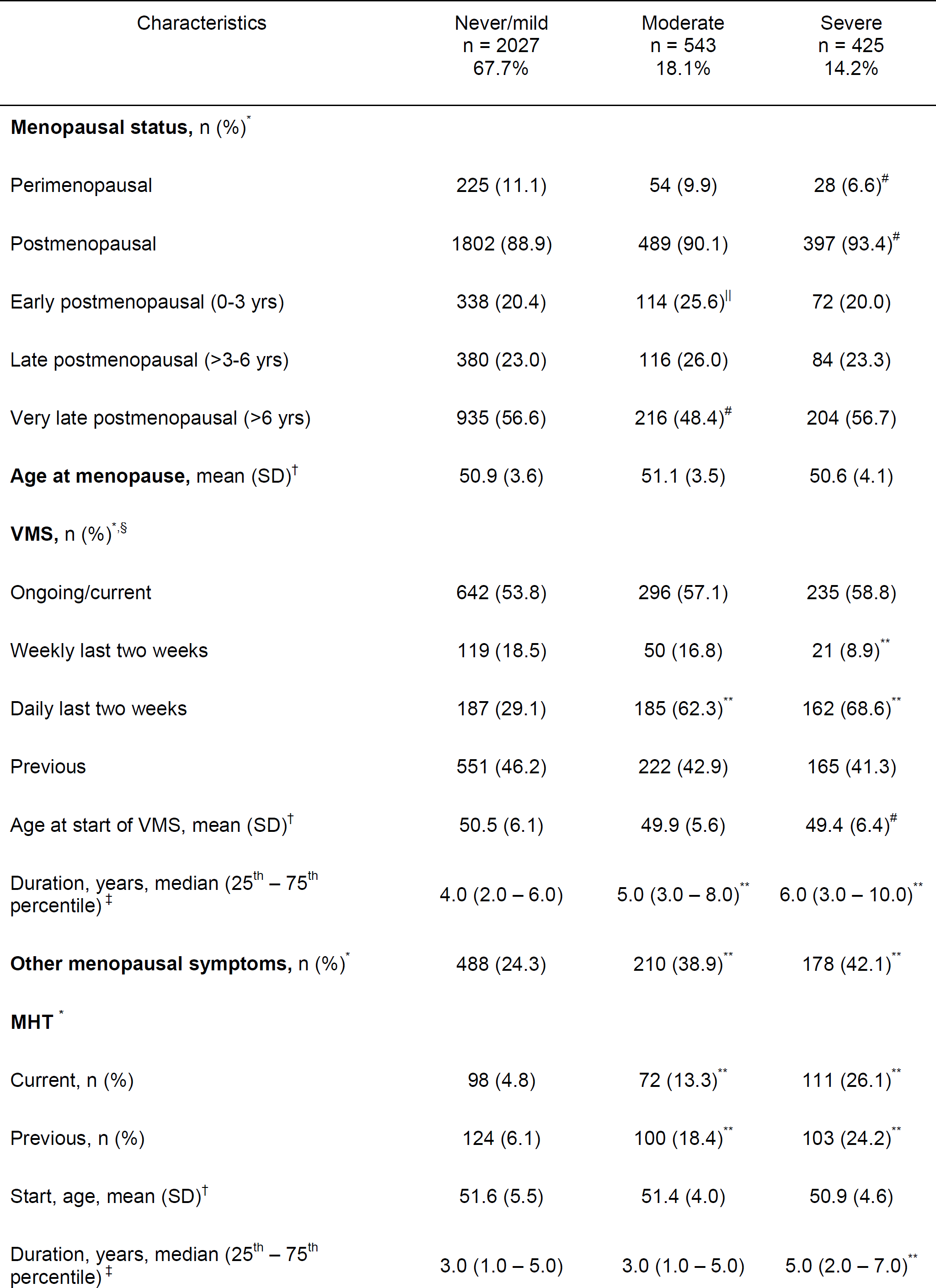
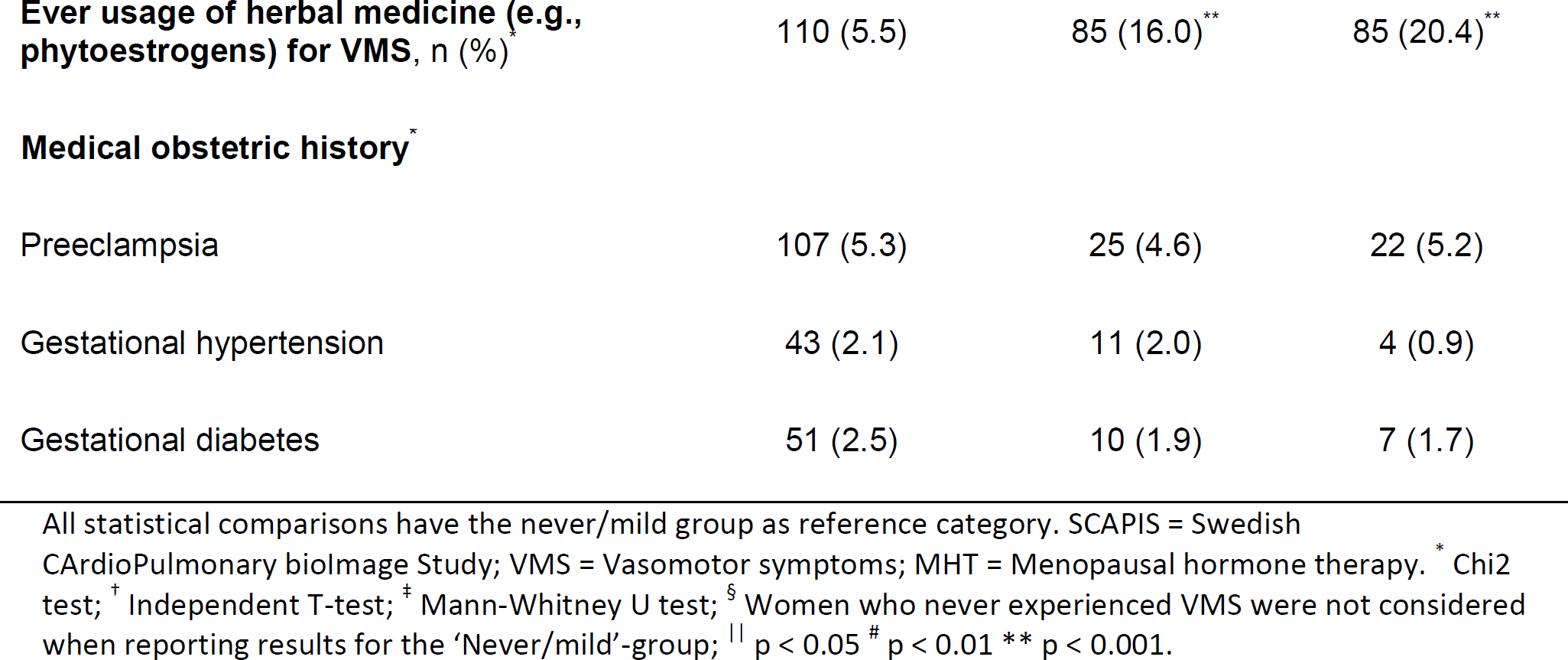
Reproduction-related characteristics by VMS category of the study population, SCAPIS (n = 2995)

| Characteristics | Never/mild<br>n = 2027<br>67.7% | Moderate<br>n = 543<br>18.1% | Severe<br>n = 425<br>14.2% |
| --- | --- | --- | --- |
| **Menopausal status, n (%)<sup>\*</sup>** |  |  |  |
| Perimenopausal | 225 (11.1) | 54 (9.9) | 28 (6.6)<sup>#</sup> |
| Postmenopausal | 1802 (88.9) | 489 (90.1) | 397 (93.4)<sup>#</sup> |
| Early postmenopausal (0-3 yrs) | 338 (20.4) | 114 (25.6) | 72 (20.0) |
| Late postmenopausal (>3-6 yrs) | 380 (23.0) | 116 (26.0) | 84 (23.3) |
| Very late postmenopausal (>6 yrs) | 935 (56.6) | 216 (48.4)<sup>#</sup> | 204 (56.7) |
| **Age at menopause, mean (SD)<sup>†</sup>** | 50.9 (3.6) | 51.1 (3.5) | 50.6 (4.1) |
| **VMS, n (%)<sup>\*,§</sup>** |  |  |  |
| Ongoing/current | 642 (53.8) | 296 (57.1) | 235 (58.8) |
| Weekly last two weeks | 119 (18.5) | 50 (16.8) | 21 (8.9)<sup>**</sup> |
| Daily last two weeks | 187 (29.1) | 185 (62.3)<sup>**</sup> | 162 (68.6)<sup>**</sup> |
| Previous | 551 (46.2) | 222 (42.9) | 165 (41.3) |
| Age at start of VMS, mean (SD)<sup>†</sup> | 50.5 (6.1) | 49.9 (5.6) | 49.4 (6.4)<sup>#</sup> |
| Duration, years, median (25<sup>th</sup> – 75<sup>th</sup> percentile)<sup>‡</sup> | 4.0 (2.0 – 6.0) | 5.0 (3.0 – 8.0)<sup>**</sup> | 6.0 (3.0 – 10.0)<sup>**</sup> |
| **Other menopausal symptoms, n (%)<sup>\*</sup>** | 488 (24.3) | 210 (38.9)<sup>**</sup> | 178 (42.1)<sup>**</sup> |
| **MHT<sup>\*</sup>** |  |  |  |
| Current, n (%) | 98 (4.8) | 72 (13.3)<sup>**</sup> | 111 (26.1)<sup>**</sup> |
| Previous, n (%) | 124 (6.1) | 100 (18.4)<sup>**</sup> | 103 (24.2)<sup>**</sup> |
| Start, age, mean (SD)<sup>†</sup> | 51.6 (5.5) | 51.4 (4.0) | 50.9 (4.6) |
| Duration, years, median (25<sup>th</sup> – 75<sup>th</sup> percentile)<sup>‡</sup> | 3.0 (1.0 – 5.0) | 3.0 (1.0 – 5.0) | 5.0 (2.0 – 7.0)<sup>**</sup> |
| **Ever usage of herbal medicine (e.g., phytoestrogens) for VMS, n (%)** | 110 (5.5) | 85 (16.0)** | 85 (20.4)** |
| **Medical obstetric history\*** |  |  |  |
| Preeclampsia | 107 (5.3) | 25 (4.6) | 22 (5.2) |
| Gestational hypertension | 43 (2.1) | 11 (2.0) | 4 (0.9) |
| Gestational diabetes | 51 (2.5) | 10 (1.9) | 7 (1.7) |

All statistical comparisons have the never/mild group as reference category. SCAPIS = Swedish CardioPulmonary bioImage Study; VMS = Vasomotor symptoms; MHT = Menopausal hormone therapy. \* Chi2 test; † Independent T-test; ‡ Mann-Whitney U test; § Women who never experienced VMS were not considered when reporting results for the 'Never/mild'-group; || p < 0.05 # p < 0.01 \*\* p < 0.001.

**Table 3.** Prevalence of subclinical ASCVD detected by CT, CCTA or ultrasound by VMS category of the study population, SCAPIS (n = 2995)

| Measures | Never/mild<br>n = 2027<br>67.7 % | Moderate<br>n = 543<br>18.1% | Severe<br>n = 425<br>14.2% |
| --- | --- | --- | --- |
| **CACS, n (%)** * |  |  |  |
| >0 | 578 (29.3) | 137 (25.7) | 143 (34.4)<sup>§</sup> |
| 1-10 | 193 (9.8) | 47 (8.8) | 51 (12.3) |
| 11-100 | 250 (12.7) | 65 (12.2) | 68 (16.3) |
| 101-400 | 107 (5.4) | 20 (3.7) | 15 (3.6) |
| > 400 | 28 (1.4) | 5 (0.9) | 9 (2.2) |
| **CACS<sup>†</sup>, median**<br>(25<sup>th</sup> – 75<sup>th</sup> percentile) <sup>†</sup> | 26.0 (6.0 – 92.0) | 22 (6.5 – 75.5) | 21 (6.0 – 71.0) |
| **CCTA, n (%)** * |  |  |  |
| Any form of coronary atherosclerosis | 516 (27.8) | 130 (25.9) | 135 (34.1)<sup>§</sup> |
| Any coronary stenosis ≥50% | 28 (1.5) | 4 (0.8) | 6 (1.5) |
| SIS >3* | 84 (4.5) | 16 (3.2) | 20 (5.1) |
| **Any carotid plaque, n (%)** * | 942 (46.6) | 255 (47.2) | 202 (47.6) |
| Unilateral plaque | 588 (29.1) | 152 (28.1) | 109 (25.7) |
| Bilateral plaques | 354 (17.5) | 103 (19.1) | 93 (21.9)<sup>§</sup> |

All statistical comparisons have the never/mild group as reference. SCAPIS = Swedish CardioPulmonary bioImage Study; VMS = Vasomotor symptoms; ASCVD = Atherosclerotic cardiovascular disease; CT = Computed tomography; CCTA = Coronary computed tomography angiography; CACS = Coronary artery calcium score; SIS = Segment involvement score. \* Chi2 test † Mann-Whitney U test; ‡ Calculation based on the participants with a CAC score >0; § p < 0.05.

VMS occurred significantly earlier in women who reported ever severe VMS, than in those who reported never/mild VMS (mean age 49.4 vs 50.5, p<0.01). The duration of VMS was significantly longer in both the severe VMS group (median 6 years) and the moderate VMS group (median 5 years) than in the never/mild group (median 4 years; p<0.01). There was a significantly higher frequency of daily VMS the past two weeks in the severe (68.6%) and moderate (62.3%) group than in the never/mild group (29.1%) (Table 2).

Among women in the study population with current or previous MHT use, 91.6% started their treatment within 6 years after menopause and the median duration of use was 4 years. Women with ever severe or moderate VMS were more likely to have used MHT compared to the never/mild group (50.3 vs 10.9% and 31.7 vs. 10.9%; p<0.01). In women with severe VMS, 24.9% had used MHT for ≥ 5 years, while the corresponding proportions in the moderate and mild VMS group were 12.2% and 3.5% (Table 2).

Women in the severe VMS group were more often born outside Sweden, had lower education, and suffered from financial strain the last year more than the never/mild VMS group. Further, they reported worse sleep habits and more continuous stress the last 5 years, were more often overweight, sedentary, past smokers and had smoked more pack years of cigarettes. Approximately 30% in this group fulfilled criteria for depression compared to 20% in the never/mild VMS group. There were no differences between VMS groups in MVPA, blood pressure or cardiometabolic biomarkers, except triglycerides, which were higher in the severe VMS group (p<0.01)(Table 1).

The prevalence of any CCTA-detected coronary atherosclerosis was higher in the severe VMS group than in the never/mild group (34.1 vs. 27.8%, p = 0.017). Similarly, CACS >0 was more prevalent in the severe VMS group than in the never/mild group (34.4 vs 29.3%, p = 0.014). For moderate VMS there were no corresponding differences compared to the never/mild group. The prevalence of SIS >3 and CACS >100 did not differ significantly between groups. The prevalence of bilateral carotid plaques was higher in the severe group compared to the never/mild group (21.9 vs 17.5%, p = 0.032), but for the moderate group the corresponding difference was not observed (19.1 vs. 17.5%, p = 0.398). The prevalence of unilateral carotid plaques did not differ significantly between groups (Table 3).

In supplemental analyses, the group who never had any VMS was compared to the group reporting mild VMS (Supplementary Tables 5-7). Women who never experienced VMS were significantly older, more often born outside Sweden, lower educated, unoccupied, and suffered from financial strain the last year more often than the group with mild VMS. They were also more frequently obese, and had on average a greater waist circumference, higher blood pressure and a worse cardiometabolic biomarker risk profile compared to women with ever mild VMS. The prevalence of ischemic stroke, hypertension, and diabetes mellitus was also higher in this group. In addition, they had higher CACS and a more widespread coronary atherosclerosis than women with mild VMS.

The proportion of missing data was <8% for the variables used in the regression models. The frequency of missing data for all variables is given in the Supplementary Table 4.

### 4.2 VMS in relation to subclinical ASCVD

The results from the multivariable regression models with subclinical ASCVD are presented in Supplementary Table 8 and illustrated in Figure 2. Severe VMS were significantly associated with coronary atherosclerosis (crude OR 1.36, 95% CI 1.08 – 1.72). The association was slightly weaker after the multivariable adjustment in model 3, OR 1.33 (95% CI 1.02-1.72) whereas additional adjustment for MHT (model 5) yielded a slightly stronger association, OR 1.41 (95% CI 1.06-1.87). In model 5, when excluding women with start of MHT > 6 years after menopause, the result remained similar (OR 1.44, 95% CI 1.08 – 1.93) (Supplementary Table 9). Moderate VMS was not significantly associated with coronary atherosclerosis in any of the models.

**Figure 2.**
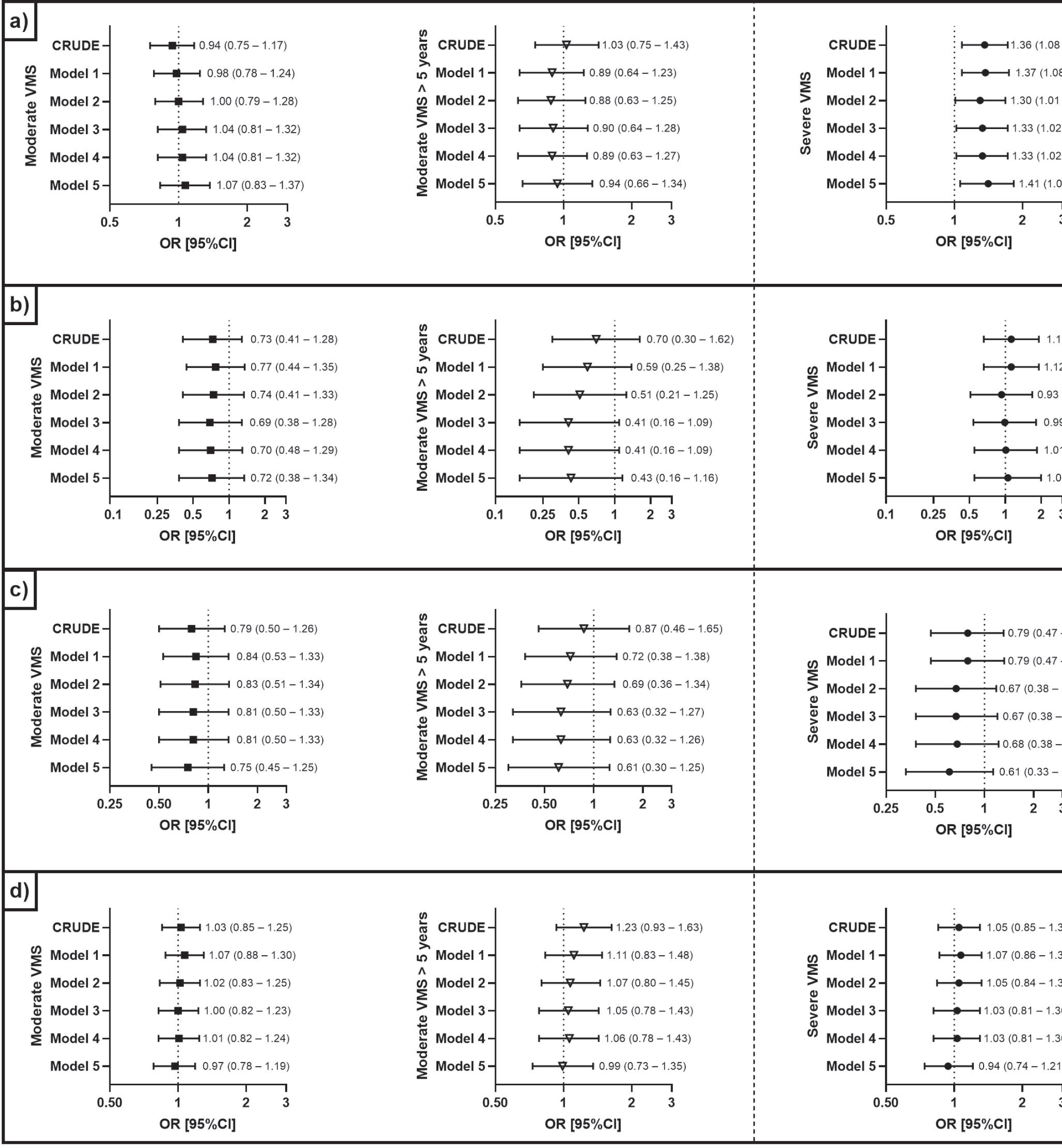
Vasomotor intensity and duration in relation to occurrence of subclinical ASCVD. The panels show results different subclinical ASCVD measures: Panel a) Any coronary atherosclerosis, Panel b) SIS >3, Panel c) CACS >100, and Panel d) any carotid plaque. All statistical comparisons have the never/mild group as reference. CRUDE: no adjustments; Model 1: age at time of study inclusion + site; Model 2: model 1 + highest degree of education, country of birth, systolic blood pressure, waist circumference, low density lipoprotein, triglycerides, diabetes mellitus, hyperlipidemia, hypertension, smoking status, and menopausal status; Model 3: model 2 + experienced stress, sleep habits, depression, and sleep apnea; Model 4: model 3 + moderate to vigorous physical activity; Model 5: model 3 + menopausal hormone therapy. SCAPIS = Swedish CArdioPulmonary bioImage Study; VMS = Vasomotor symptoms. SCAPIS = Swedish CArdioPulmonary bioImage Study; ASCVD = Atherosclerotic cardiovascular disease; VMS = Vasomotor symptoms; Segmental involvement score; CACS = Coronary artery calcium score.

If severe VMS lasted >5 years, the association with coronary atherosclerosis was more pronounced, crude OR 1.70, 95% CI 1.26 – 2.30 (Figure 2 and Supplementary Table 10). This association was weakened but still significant in model 1, (OR 1.48, 95% CI 1.09 – 2.01), and similar in model 2 (OR 1.49, 95% CI 1.07 – 1.97), model 3, (OR 1.53, 95% CI 1.09 – 2.14) and model 4 (OR 1.50, 95% CI 1.07 – 2.11). In model 5, where MHT was added, the association was strengthened (OR 1.67, 95% CI 1.16 – 2.40) (Figure 2). For severe VMS with shorter duration (≤5 years), the association with coronary atherosclerosis was not significant (Supplementary Table 10). For moderate VMS, no effect of symptom duration was observed in relation to coronary atherosclerosis (Supplementary Table 10). For severe VMS that started before menopause, but not for VMS that started after menopause, a significant association with coronary atherosclerosis was observed that persisted after multivariable adjustments, model 3 OR 1.60 (95% CI 1.06-2.42) and model 5 OR 1.69 (95% CI 1.10 - 2.60) (Supplementary Table 11).

Neither moderate nor severe VMS were significantly associated with CACS >100, widespread coronary disease (SIS >3) or any carotid plaque (Supplementary Table 8, Figure 2). In supplementary analyses, severe VMS were significantly associated with CACS >0 in the crude model but not after multivariable adjustments (Supplementary Table 12). Severe VMS were significantly associated with bilateral carotid plaques in crude analyses and borderline significant after multivariable adjustments (Supplementary Table 13). For unilateral plaque, no significant association was observed.

## 5. Discussion

In this nation-wide population-based study, we found that ongoing or previous severe, but not moderate, VMS were significantly associated with coronary atherosclerosis detected with CCTA, independent of a wide range of lifestyle, socioeconomic and clinical cardiovascular risk factors. To our knowledge this is the first time CCTA data have been used assessing the association between VMS and subclinical ASCVD.

In keeping with a previous study (13), women who experienced severe VMS had the earliest onset and longest duration of VMS. Arguably, VMS duration could have bearing on the strength of the association to coronary atherosclerosis. To the best of our knowledge, this has not been studied previously, and interestingly, our results show that severe VMS lasting >5 years were significantly associated with coronary atherosclerosis, whereas no corresponding association was observed for shorter durations. Moderate VMS, however, was not associated with coronary atherosclerosis regardless duration. We also found that pre-menopausal but not post-menopausal onset of severe VMS was associated with coronary atherosclerosis. These findings agree with the American Study of Women’s Health Across the Nation, which reported associations between frequent VMS persistence over time and incident CVD (25).

The findings are also consistent with results from the Womeńs Ischemia Syndrome Evaluation (WISE) that show associations between early onset VMS and reduced endothelial function and CVD mortality (35). However, results from a study that pooled data from six different prospective studies show that both pre-menopausal and post-menopausal onset of VMS are associated with incident CVD (26).

The persisting association between severe VMS and coronary atherosclerosis after adjustment for conditions known to be prevalent in women with VMS, such as sleep disturbances and depression (13, 14), is on the one hand surprising as VMS could have a negative impact on lifestyle and sleep quality, which in turn could increase the risk of ASCVD. On the other hand, our results are consistent with several previous studies of VMS in relation to subclinical CVD and incident manifest CVD where no substantial impact on the results were observed when adjusting for such covariates (22–26). Although it cannot be ruled out that severe VMS is merely a marker of an unfavorable cardiovascular risk profile, our findings of an independent relationship between severe VMS and coronary atherosclerosis could indicate an unknown underlying pathophysiological mechanism, either in the initiation or the progression of the atherosclerotic process in midlife women. Further research is required to elucidate mechanisms by which VMS may exert a deleterious effect on the coronary arteries, but we can speculate that women with long-term severe VMS or whose severe VMS started early may have increased sensitivity to hormonal fluctuations, which in turn may contribute to atherosclerotic development via autonomic neurovascular dysregulation (36). Thus, these women may in general have an underlying adverse vascular reactivity manifested during the menopausal transition. Effects could perhaps be related to an over-reactivity of the sympathetic nervous system, with inhibition of cardiac vagal control (37), or to changes in the hypothalamic-pituitary-adrenal (HPA) axis associated with variable cortisol concentrations (38, 39). Furthermore, because the fast withdrawal of estrogen during menopausal transition rather than the absolute plasma level may be connected to an instability in the thermoregulatory center in hypothalamus (40), it seems reasonable that increased central thermoregulatory sensitivity or hampered adaptation capacity may indirectly be related to a heightened central sympathetic tone, resulting in an over-activity of the sympathetic nervous system (40, 41).

The analyses of VMS in relation to the other subclinical ASCVD measurements – CACS >100, SIS >3 and any carotid plaque - showed no significant associations. To improve comparison to previous study results, we added analyses on VMS in relation to CACS >0. Still, we observed no significant associations after multiple adjustments (Supplementary Table 12). These findings are in line with most previous studies on VMS in relation to CACS and/or carotid atherosclerosis(21), with some exceptions (22, 23). CCTA is a diagnostical and non-invasive angiographic method able to detect coronary artery disease (CAD), compared to CAC that rather predicts risk for future manifest CVD events (42). While CACS reflects the sum of coronary calcifications, CCTA can detect calcified as well as non-calcified plaques, degree of plaque stenosis, and extended plaque features such as plaque stability and anatomical distribution (43). Thus, VMS might be associated to early coronary atherosclerosis, already before the development of calcified plaques, which might explain the observation of a strong and independent association for CCTA-detected coronary atherosclerosis but not with coronary calcification. Since SIS >3 was uncommon in the included age group, we might lack statistical power to detect significant differences and thus cannot rule out true associations. Although the atherogenesis appears to be similar for both coronary and carotid plaques (44), the lack of significant associations between severe VMS and any carotid plaque could relate to a different pathophysiology behind the development of coronary and carotid atherosclerosis. However, in the supplementary analyses severe VMS were borderline significantly associated with bilateral carotid plaque, which compared to unilateral carotid disease has been shown so be a better predictor of the 5-year risk of coronary death (45). Although this may be a spurious finding, there is a possibility that unilateral carotid atherosclerosis is rather caused by anatomical and hemodynamic factors, while bilateral carotid plaques reflect a more extensive atherosclerotic disease with systemic factors giving rise to its development (45).

For analyzes of VMS in relation to subclinical ASCVD, the choice of “reference group” is not obvious and varying definitions of “bothersome” symptoms makes comparisons between studies challenging. In previous studies, women without VMS have usually constituted the reference group. We found that women who never experienced VMS had significantly more cardiovascular risk factors and coronary atherosclerosis than women with mild VMS, a finding consistent with the results from the WISE study, which observed an increased CVD mortality in this group compared with women with late-onset VMS (35). Thus, women without VMS may not always have the lowest risk of developing ASCVD and thus our reference category also included women who reported mild VMS, which constituted the largest group in our study population (43.3%) (Supplementary Table 14).

Women with severe VMS performed MVPA to a similar extent as women with no or mild VMS. Neither did self-reported level of physical activity differ significantly between the groups. Hence, we found no indication that physical activity would mitigate a potential detrimental effect of VMS on the coronary arteries. This somewhat contradicts previous results showing that physically active as compared to sedentary postmenopausal women report less VMS (20). Although the current study was not designed to study treatment effects, it is relevant to note that the association between >5 years of VMS and coronary atherosclerosis became stronger when adjusting for MHT, as this could possibly signal a mitigating effect. However, this result must be interpreted with caution due to bias related to confounding by indication. Contraindications for prescribing MHT have since the WHI-study (46) included a history of cardiovascular disease, which means a selection of cardiovascular healthier women prescribed MHT. On the other hand, VMS severity could also influence whether the woman is treated with MHT or not, which, under the assumption that severe VMS affect the development of subclinical atherosclerosis, means a selection against cardiovascular less healthy women prescribed MHT.

There are several limitations to our study. Firstly, the cross-sectional observational study design limits any causal conclusions to be drawn since we do not know when VMS appeared in relation to the atherosclerosis development. Secondly, the results may not be generalizable to other geographical regions, ethnicities and age-groups since the definitions and prevalence of VMS differ (10, 13). Thirdly, the assessment of VMS may be subject to recall bias, given that up to 10-15 years could have passed since the study participants experienced their symptoms. However, it is likely that such recall bias would be non-differential because the women were not aware of their measures of subclinical coronary atherosclerosis or other cardiovascular risk assessments when they completed the questionnaire. In that case, this would have a weakening effect on our results.

## 6. Conclusions

Ongoing or previous severe VMS, with >5 years duration or beginning before menopause, were significantly associated with subclinical CCTA-detected coronary atherosclerosis, independent of a wide range of socioeconomic, demographic, lifestyle-related and clinical cardiovascular risk factors. No corresponding associations were observed for SIS >3, CACS >100 or any carotid plaque. Further research is required to establish a possible causal relationship between severe VMS and coronary atherosclerosis, as well as the pathophysiological mechanism behind such association. However, our findings support a more consistent implementation of preventive cardiovascular strategies in gynecological care, targeting women with severe VMS which may be a marker of poor or ongoing deterioration of cardiovascular health. Here is a window of opportunity for preventing cardiovascular disease in midlife women, as severe VMS increase the propensity to seek medical advice.

## Data Availability

The authors declare that all supporting data are available within the article.

## Acknowledgements

We would like to thank Professor and MD Mats Hammar for his support in conceptualization of the SWH questionnaire. In addition, authors would like to thank Max Vikström (Karolinska Institutet, Unit of Cardiovascular and Nutritional Epidemiology, Institute of Environmental Medicine, Stockholm, Sweden) for his assistance and support with the statistical analyses.

## Author contributions

Sigrid Nilsson contributed to investigation, validation, visualisation, methodology, formal analysis, writing the original draft, reviewing, and editing the manuscript.

Angelika Qvick contributed to investigation, validation, visualisation, methodology, writing the original draft, reviewing, and editing the manuscript.

Moa Henriksson contributed to data curation, reviewing, and editing the manuscript.

Anna-Clara Spetz Holm contributed to supervision, conceptualisation, project administration, validation, investigation, reviewing and editing the manuscript.

Sofia Sederholm Lawesson contributed to supervision, validation, investigation, methodology, reviewing and editing the manuscript.

Karin Leander contributed to supervision, conceptualisation, project administration, data curation, methodology, validation, investigation, reviewing and editing the manuscript, and funding acquisition.

## Abbreviations

ASCVD: – atherosclerotic cardiovascular disease
CAC: – coronary artery calcium
CACS: – coronary artery calcium score
CCTA: – coronary computed tomography angiography
C-IMT: – carotid intima media thickness
CT: – computed tomography
CVD: – cardiovascular disease
MHT: – menopausal hormone therapy
MVPA: – moderate to vigorous physical activity
SCAPIS: – Swedish CArdioPulmonary bioImage Study
SIS: – segment involvement score
SWH: – Study of Women’s health
VMS: – vasomotor symptoms

## Clinical Perspective

- What is new?

° Previous or ongoing severe vasomotor symptoms were, independent of a wide range of cardiovascular risk factors, significantly associated with coronary atherosclerosis identified by coronary computed tomography angiography screening of the general population, aged 50-65 years.
° The association was limited to women with a duration of severe vasomotor symptoms of more than five years or whose severe vasomotor symptoms began before menopause onset.
- What are the clinical implications?

° Our results reflect a need for extra vigilance regarding cardiovascular risk factors in women suffering from severe vasomotor symptoms.

### Supplemental Material

Supplemental methods Supplemental tables 1-14

